# Leveraging Artificial Intelligence and Data Science for Integration of Social Determinants of Health in Emergency Medicine: A Scoping Review

**DOI:** 10.1101/2023.10.17.23297158

**Authors:** Donald Apakama, Ethan E Abbott, Lynne Richardson, Lili Chan, Brendan G Carr, Girish N Nadkarni

## Abstract

**Objective:** Social Determinants of Health (SDOH) are critical drivers of health disparities and patient outcomes. However, accessing and collecting patient level SDOH data can be operationally challenging in the emergency department clinical setting requiring innovative approaches. This scoping review examines the potential of artificial intelligence (AI) and data science for modeling, extraction, and incorporation of SDOH data specifically within emergency departments (ED), further identifying areas for advancement and investigation.

**Methods:** We conducted a standardized search across Medline (Ovid), Embase (Ovid), CINAHL, Web of Science, and ERIC databases for studies published between 2015-2022. We focused on identifying studies employing AI or data science related to SDOH within emergency care contexts or conditions. Two specialized reviewers in Emergency Medicine and clinical informatics independently assessed each article, resolving discrepancies through iterative reviews and discussion. We then extracted data covering study details, methodologies, patient demographics, care settings, and principal outcomes.

**Results:** Of the 1,047 studies screened, 26 met the inclusion criteria. Notably, 9 out of 26 studies were solely concentrated on ED patients. Conditions studied spanned broad Emergency Medicine complaints and conditions including sepsis, acute myocardial infarction, and asthma. The majority (n=16) explored multiple SDOH domains, with homelessness/housing insecurity and neighborhood/built environment predominating. Machine learning (ML) techniques were utilized in 23 of the studies, natural language processing (NLP) being the most common approach used (n=11). Rule-based (n=5), deep learning (n=2), and pattern matching (n=4) were the most common NLP techniques used. NLP models in the reviewed studies displayed significant predictive performance with outcomes, With F1-scores ranging between 0.40 - 0.75 and specificities nearing 95.9%.

**Conclusion:** Although in its infancy, the convergence of AI and data science techniques, especially ML and NLP, with SDOH in Emergency Medicine offers transformative possibilities for better usage and integration of social data into clinical care and research. With a significant focus on the ED and notable NLP model performance, there is an imperative to standardize SDOH data collection, refine algorithms for diverse patient groups, and champion interdisciplinary synergies. These efforts aim to harness SDOH data optimally, enhancing patient care and mitigating health disparities. Our research underscores the vital need for continued investigation in this domain.

## Introduction

Medical care, while crucial, contributes to only about 20% of the modifiable factors influencing a population’s health outcomes, while 80% are influenced by genetics, individual behaviors, and socioeconomic factors. The latter two form the Social Determinants of Health (SDOH)^1^ that operate at various levels. From macroeconomic policies of nations to public education and housing policies, these structural factors shape resource distribution and societal positions. Consequently, they influence living conditions, access to essential resources, and daily life circumstances, ultimately molding health, and health disparities.^2^ Every patient’s health trajectory is influenced by SDOH, which can manifest positively (e.g., high income, food security) or adversely.^3^ The negative aspects can be categorized into social risks, conditions linked to poor health, and social needs, which are individual preferences for assistance.^4^ These determinants, especially when adverse, can hinder optimal care and impact clinical outcomes.^5^

Emergency Medicine (EM) is a unique medical specialty. It can both identify and address adverse SDOH, making it a pivotal setting for intervention. The high prevalence of social needs among emergency department (ED) patients, especially those with low socioeconomic status, housing insecurity, or limited access to care, underscores the potential of ED-based SDOH interventions.^6, 7^ However, there are significant challenges; comprehensive social risk screenings in the ED are often impractical due to patient volume, acuity, and health system financial constraints. Relying solely on electronic health records (EHRs) is time-consuming and inconsistent. Furthermore, the scattered and unstructured nature of SDOH data in EHRs makes it difficult for ED physicians to identify patients with adverse SDOH.^8–10^

Social informatics refers to the interdisciplinary study of the design, uses, and consequences of information technologies in the context of social and organizational settings. It bridges the gap between the technical and social worlds, offering insights into how technology and societal factors interplay. The vast possibilities of social informatics lie in its potential to reshape how we understand, interpret, and act on social data in healthcare settings. By integrating social data with health data, it aims to enhance clinical care and overall health outcomes.^11^ Techniques like Natural Language Processing (NLP), Artificial Intelligence (AI), and Machine Learning (ML) are being harnessed to extract, utilize, and model SDOH data effectively.^12–14^ While existing literature has touched upon SDOH in the ED, a comprehensive review focusing on the application of data sciences in this context is lacking. This scoping review aims to map the current literature, pinpoint areas for future research, and highlight the transformative potential of integrating data sciences into EM SDOH research.

## Methods

### Data Sources and Literature Search Strategy

To capture the evolving role of AI data science in Emergency Medicine, particularly around a SDOH and social context, we searched the literature from 2015 to 2022, a period marked by rapid advancements in AI applications in healthcare. We included articles from databases such as Medline (Ovid), Embase (Ovid), CINAHL, Web of Science, and ERIC, prioritizing research that melded data science with emergency care settings.

Our search encompassed terms related to SDOH, data science techniques such as machine learning algorithms, natural language processing, and artificial intelligence, and Emergency Medicine (see supplemental table 1 for search terms utilized).

### Article Selection

We focused our review on studies leveraging data science techniques to extract or model SDOH data in EM. Recognizing the paucity of EM-specific research using AI/ML algorithms, we also considered studies on emergency-related conditions that might be seen in other clinical non-ED settings. This included conditions such as opioid use disorder (OUD), HIV, and epilepsy. We intentionally excluded COVID-19 studies, given their unique characteristics and volume of literature. This exclusion ensured a more focused review with current relevance. Two independent reviewers (DA and EA) assessed titles and abstracts for final inclusion. Any disagreements were resolved through joint discussions. We utilized Covidence (Covidence systematic review software, Veritas Health Innovation, Melbourne, Australia), a standardized systematic review software.

### Data Extraction

We extracted data from the selected studies using a standardized form, ensuring uniformity. We captured study objectives, methods, clinical care setting, machine learning algorithms, modeling approaches, and specified outcomes (see Supplemental Table 2). We focused on data science, AI and machine learning algorithms, SDOH domains, and clinical outcomes. While our focus remains descriptive, we abstained from a quality assessment, aligning with standard scoping review guidelines.

## Results

### Overall Study Characteristics

After screening 1,047 studies, 26 met our final inclusion criteria (Figure 1 and Supplemental Table 1). We excluded a significant number of studies because they did not focus on Emergency Medicine conditions or complaints and did not employ AI/ML techniques in overall approach to study question. Most studies were published after 2020 (Figure 2) and included patient populations focusing exclusively on the ED (n=9), pediatric patients (n=2), patients treated by emergency medical services (EMS) (n=2), and US Veterans (n=2) among examples.

**Figure 1:**
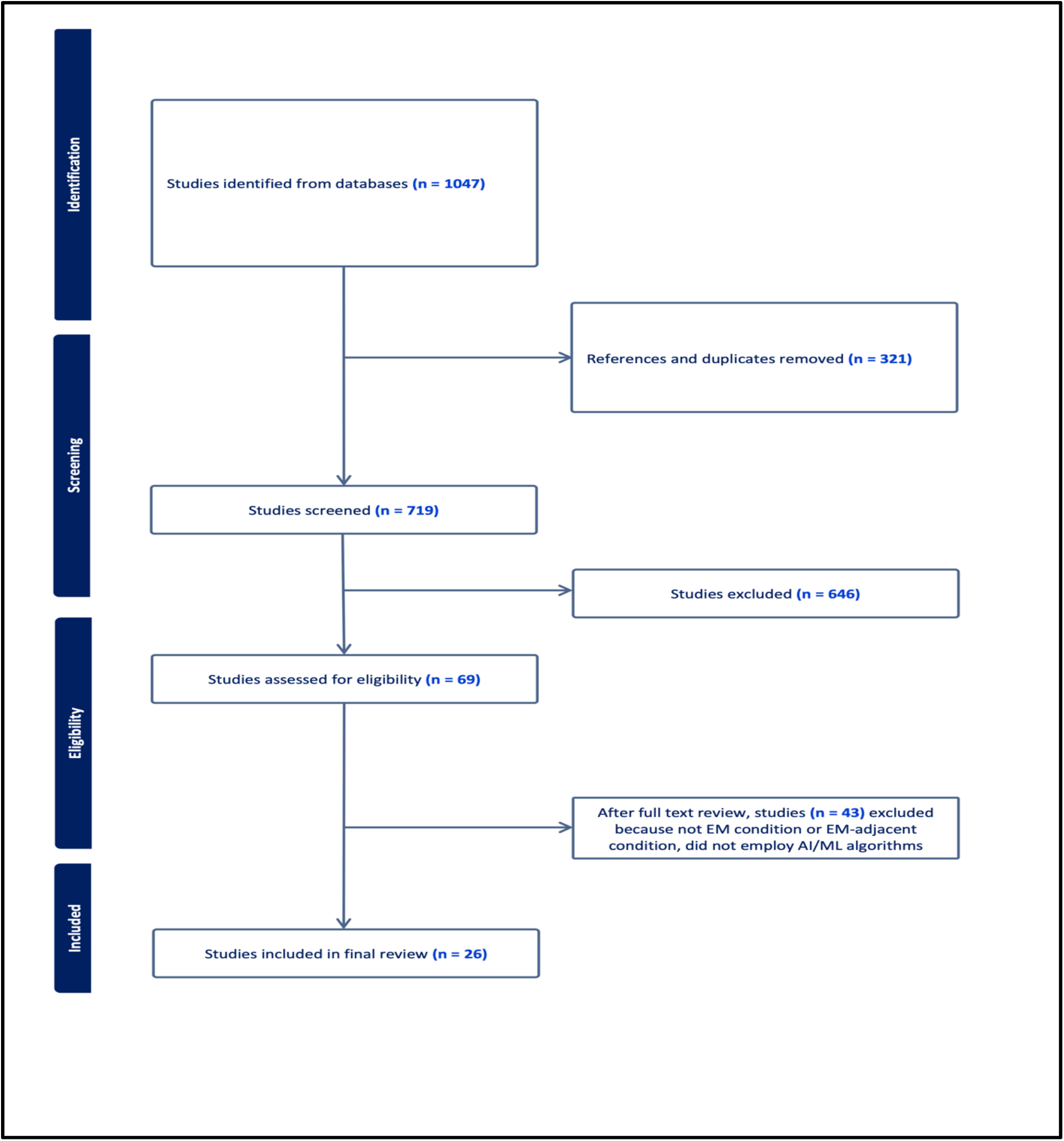
Preferred Reporting for Systematic Reviews and Meta-Analysis Flow Chart (PRISMA) Diagram. 2015-2022 search of Medline (Ovid), Embase (Ovid), Cumulative Index to Nursing and Allied Health Literature (CINAHL), Web of Science, and Education Resource Information Center (ERIC).

**Figure 2:**
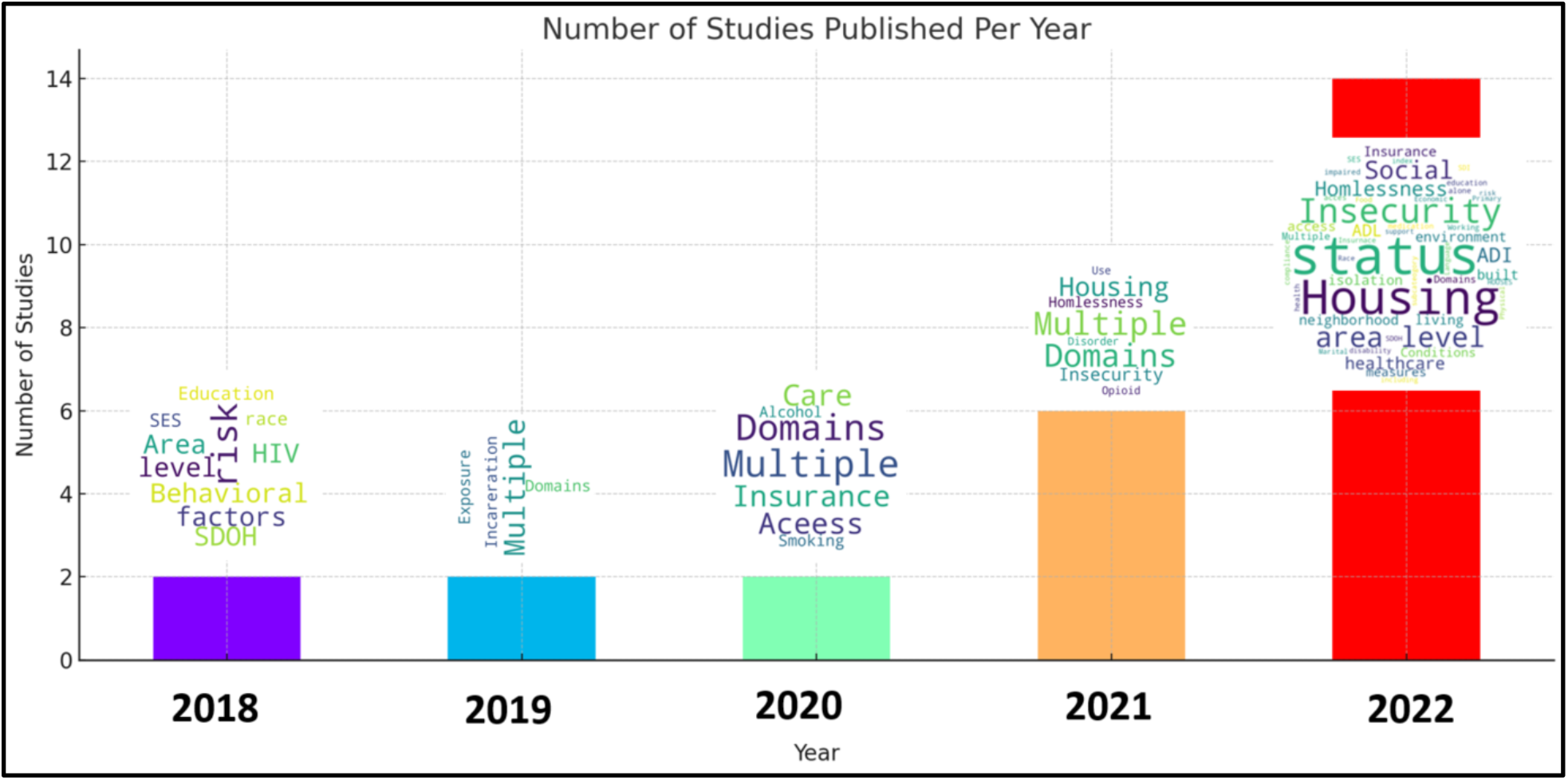
Number of publications by year (2015-2022) identified in this review. Word clouds show main themes of papers for the corresponding year.

### SDOH Domains

Over 60% of studies we identified (n =16) employed, modeled, or extracted features across multiple SDOH domains, resulting in significant overlap across publications included in this review (figure 2 word cloud). These domains included housing insecurity and homelessness, neighborhood and built environment, income and socioeconomic status, employment, family and social support, food insecurity, insurance status and stability, and history of incarceration. While individual level SDOH data was the prevalent unit of analysis, 5 studies used area level data or aggregated measures such as the Social Deprivation Index (SDI), Area Deprivation Index (ADI), or the Gini coefficient. Housing insecurity and homelessness emerged as the most predominate SDOH domains assessed among 23% of the studies identified (n=6). The domain of neighborhood and built environment was also present across multiple studies and the focus of several publications (n=4). Exposure to or history of incarceration (n=2) as well as opioid use disorder (n=2) were also notable.

### Exploration of Emergency Medicine Conditions

The scope of EM clinical conditions and complaints that studied were broad, including sepsis, acute myocardial infarction, heart failure, asthma, diabetes, chest pain, and epilepsy (Figure 3). Sepsis was the only specific EM condition we identified in more than one study (n =2). Several studies focused on all cause ED revisits (n=2), “preventable visits” and admissions (n=2), and ED utilization (n=2).

**Figure 3:**
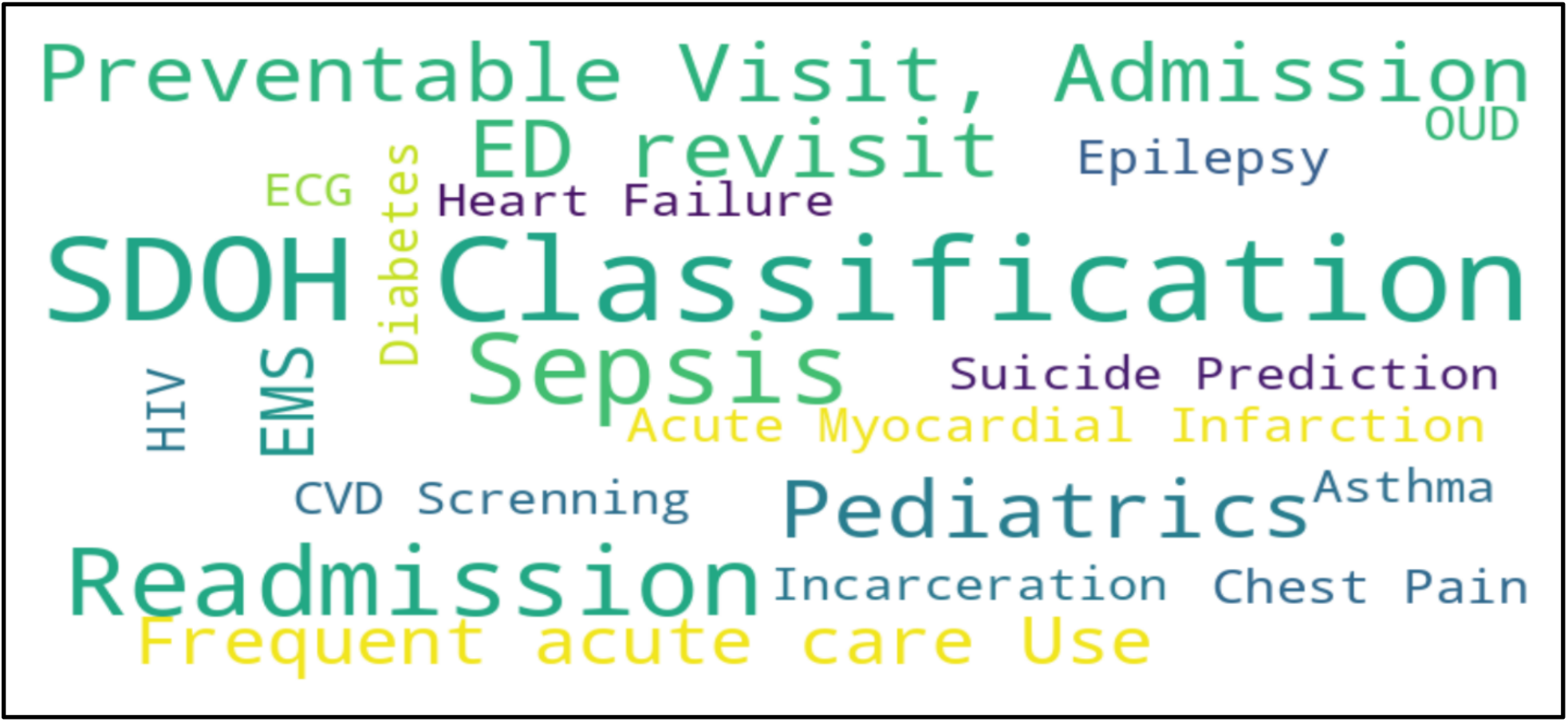
Emergency Medicine Conditions Word Cloud.

### AI/ML Algorithms

Machine learning (ML) techniques were employed in 23 studies, encompassing methods like random forest, CART, support vector machines, neural networks, and natural language processing (NLP) (Figures 4 and 5). Of these, Random Forest emerged as the most popular (n = 13), closely followed by NLP (n =11). Key algorithms are discussed in in further detail.

**Figure 4:**
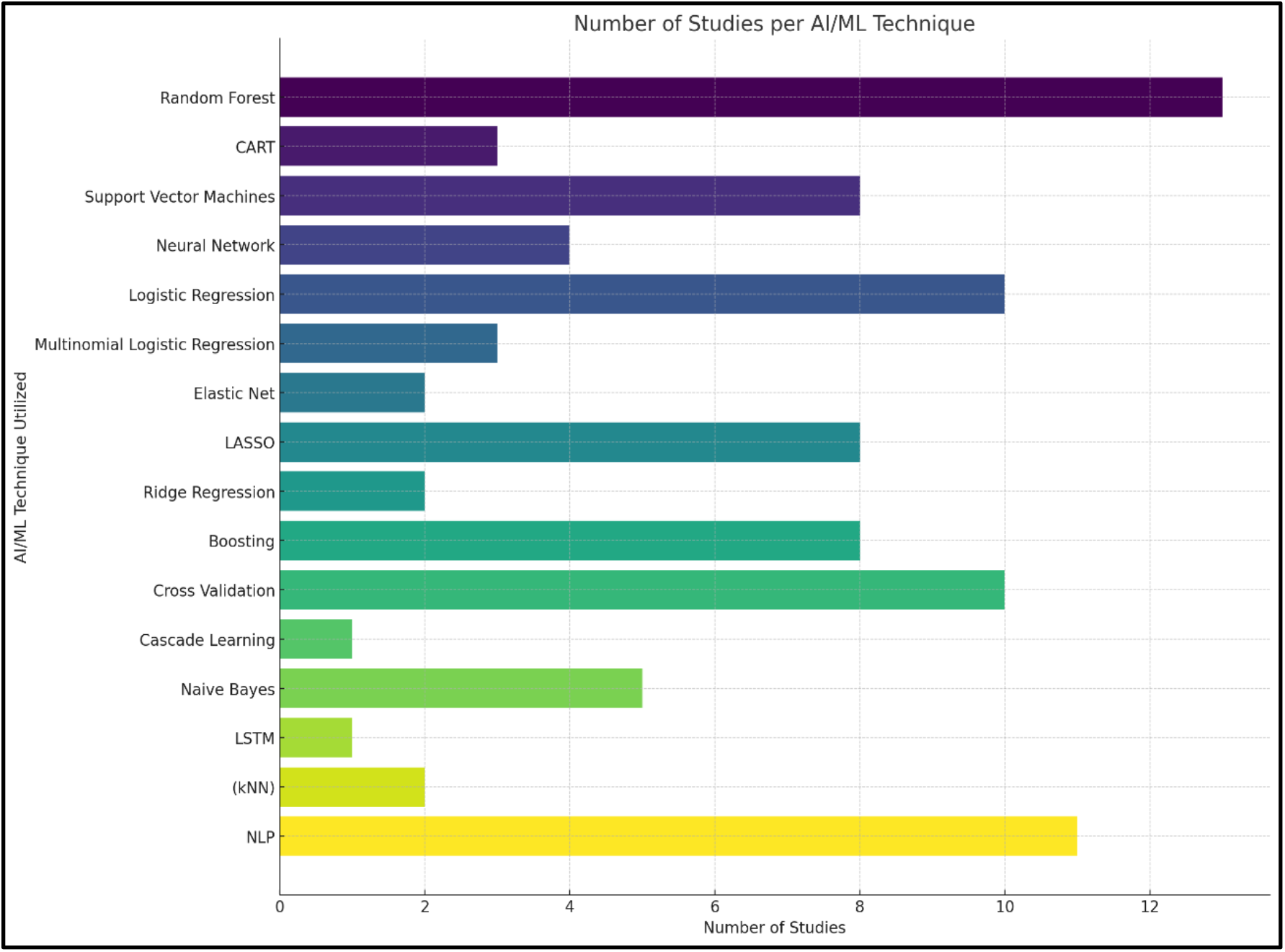
Overall counts of AI/ML algorithms employed.

**Figure 5:**
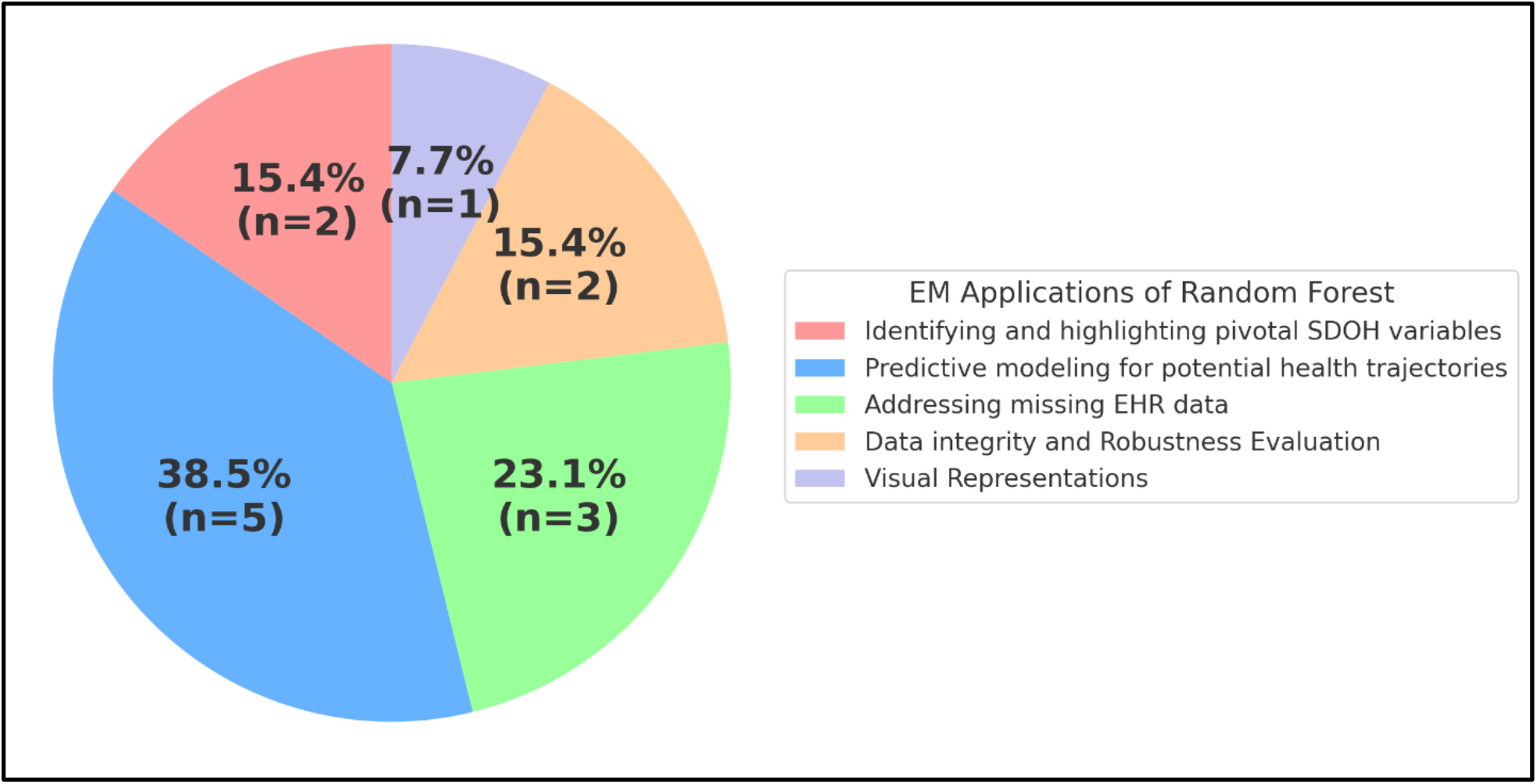
Overall counts/percentages of Random Forest techniques employed for extraction of SDOH data.

### Random Forest in SDOH Variable Classification and Data Management

Random Forest, an advanced ensemble machine learning method, was notably present across multiple studies identified in this review. This technique was used to discern and highlight pivotal SDOH variables. Its ability to create predictive models, offering foresight into potential health trajectories based on the subtleties of SDOH indicators, was also evident. The most common use of Random Forest, however, was to address missing EHR data, ensuring the integrity and robustness of analyses. Beyond its analytical capabilities, it was also used to create insightful visual representations, offering a comprehensive view into the intricate web of variables, their interactions, and their overarching impact on disease states like ACS, Epilepsy, Asthma, and OUD.

### Natural Language Processing for SDOH Data Extraction

The integration of Natural Language Processing (NLP) in the realm of Emergency Medicine offers a promising avenue for the precise extraction of Social Determinants of Health (SDOH) from electronic health records (EHR). Traditional methodologies, such as manual reviews and rudimentary keyword searches, are increasingly recognized for their inherent limitations, particularly in the context of vast and intricate EHR datasets. Our scoping review elucidated the prevalence of several NLP techniques in the field. Text representation methods like Term Frequency - Inverse Document Frequency (TF-IDF) (n = 4), Bag of Words (n = 3), and Word2Vec (n = 1) were prominently utilized, underscoring their fundamental role in converting textual data into computationally amenable formats (figure 6). In the realm of topic modeling and semantic analysis, Latent Dirichlet Allocation (LDA) was noted in 2 studies, highlighting its potential in discerning latent topics within medical records. Approaches favored Rule-Based methodologies, found in 5 studies, followed by Deep Learning (n = 2) particularly structures like BI-LSTM (Bi-directional long short-term memory) and Pattern Matching (n = 4). From a software perspective, both proprietary (n = 5) and open source (n = 6) tools were harnessed, reflecting the diverse ecosystem of NLP tools available for research. These NLP methodologies, especially the dominant ones like Bag of Words, TF-IDF, and deep learning structures, have demonstrated notable efficacy. However, to achieve the pinnacle of precision in SDOH data extraction, it is imperative to continually refine these NLP techniques. Collaborative endeavors involving domain experts, iterative model training, and the assimilation of multifaceted data sources are paramount to enhancing the accuracy and relevance of extracted insights.

**Figure 6:**
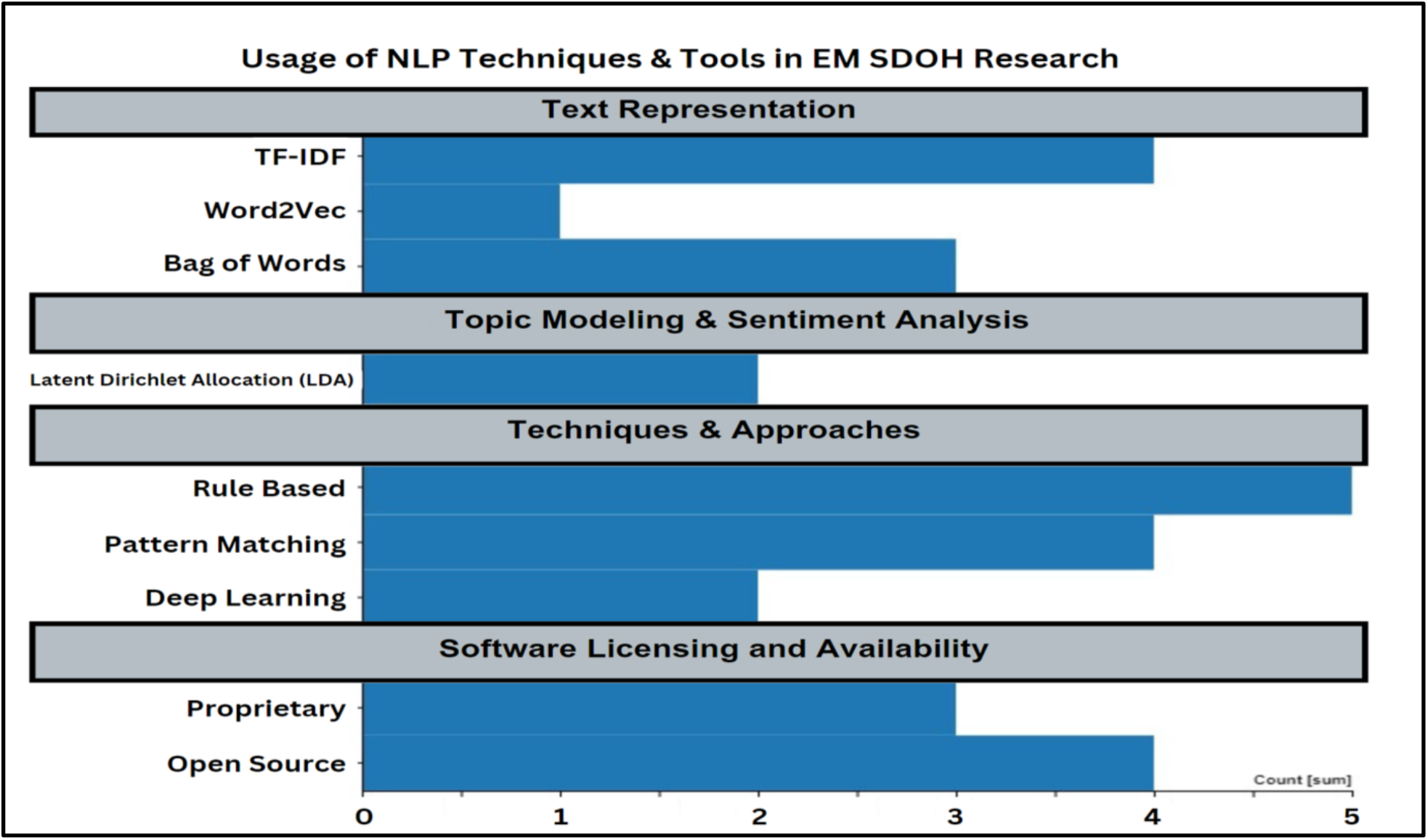
Overall counts of NLP techniques employed for extraction of SDOH data.

### Health Information Exchange (HIE) for SDOH Data Aggregation

HIE, featured in 4 out of 26 studies, aggregates patient data across healthcare entities, providing a comprehensive view of a patient’s clinical journey. With 40% of patients in one study having encounters at multiple organizations, the importance of HIE in reflecting the distributed nature of ED care becomes evident. HIE can aid in analyzing care transitions and augmenting the sample size and diversity for SDOH research. However, challenges like data sharing, data quality, and privacy regulations need to be addressed. In essence, HIE holds immense potential for Emergency Medicine research, offering both multi-organizational and community-level insights.

**Figure 7:**
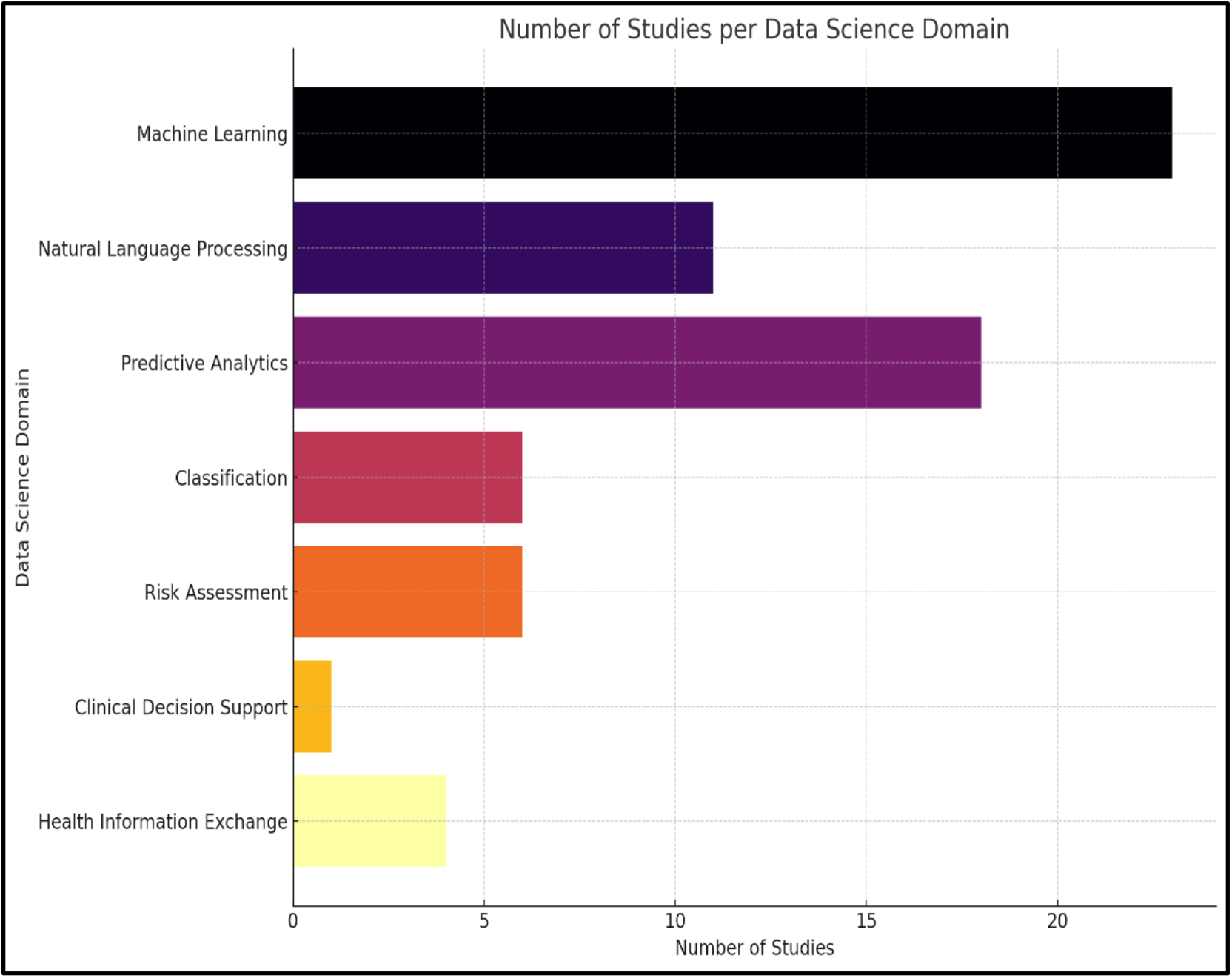
Overall counts for identified articles by AI/ML Domain and/or task.

### Predictive Applications

A total of 19 studies presented a diverse array of machine learning strategies, each tailored for distinct predictive outcomes. The efficacy of predictive models was a pivotal outcome in 10 studies. Performance metrics for these models, when used as a primary outcome, showcased a range of F1-scores from 0.4 up to 0.75, indicating varied precision and recall across studies. These ranged from supervised models like random forest classifiers for acute coronary syndrome predictions to neural networks targeting sepsis-related readmission risks. Notably, ensemble methods were adeptly utilized to discern primary risk factors for opioid use disorders. Within this realm, NLP proved instrumental, particularly in classification tasks, risk stratification, and fortifying clinical decision-making processes. Beyond this, key outcomes highlighted patterns in ED revisits (n = 6), in-hospital mortality (n = 2), algorithmic bias (n =1), and mismatches between physician annotations and claims data (n =1).

### Data Quality, Privacy, and Algorithmic Bias

In the comprehensive spectrum of studies analyzed, an intriguing observation surfaced: a noticeable gap in the examination of data quality and privacy concerns within the realm of SDOH and Emergency Medicine. Despite sifting through a multitude of research articles, we did not encounter any study that directly tackled these pivotal issues. This omission underlines the potential vulnerabilities in the application of AI and ML techniques, especially when handling sensitive patient data. Furthermore, a singular study broached the topic of algorithmic bias, a topic of paramount importance given the potential repercussions on healthcare outcomes and equity. The underrepresentation of these themes hints at uncharted territories that warrant meticulous exploration in future research endeavors, ensuring that the integration of AI and ML in Emergency Medicine is both robust and ethically sound.

## Discussion

The leveraging of SDOH data within EM research and clinical care is pivotal for gaining new insights, improving patient outcomes, and optimizing healthcare delivery. Our scoping review highlights the transformative role of data science, chiefly AI/ML, and the subdiscipline of NLP, in improving SDOH data integration and modeling within EM. Emerging from our review is an increase towards employing data science to harness, operationalize, and model SDOH in emergency care settings. This progression signifies a shift: a pivot towards a comprehensive, data-infused approach that addresses not just emergent conditions but also the intricate web of social and economic determinants impacting health. NLP excels at extracting SDOH information from the unstructured text of electronic health records, while ML’s predictive strength can transform these insights into actionable predictions. Such models, equipped with SDOH data, can catalyze precision interventions, potentially identifying mechanisms for ED revisits, in-hospital mortality, and readmissions.

Delving deeper into the SDOH domains, housing insecurity and the neighborhood environment emerged as primary determinants, witnessing significant attention across the studies. Their frequent appearance in the research landscape underscores their profound impact on health outcomes within emergency settings. While these domains were at the forefront, other determinants like education, employment, and social networks were also featured, albeit to a lesser extent. The emphasis on these SDOH domains, especially housing insecurity, suggests a pressing need for targeted interventions and policies within emergency care settings. As the healthcare sector continues to evolve, understanding these predominant SDOH domains and harnessing the power of data science will be pivotal in offering a more holistic, patient-centric approach to emergency care.

Our comprehensive review, while offering insights, bears certain limitations warranting acknowledgment. The time frame for our study, confined to 2015 to 2022, captures most contemporary advancements but might inadvertently omit foundational studies predating this period, potentially offering evolutionary insights. While we highlighted NLP and other ML techniques, the vast expanse of data science boasts other emerging tools such as the recent and rapid development of Large Language Modeling (LLMs) such as ChatGPT, which were absent in our review. The potential applications of LLMs in harnessing and modeling SDOH within the EM setting are rapidly emerging and will likely expand the possibilities for improving health outcomes and disparities.

Secondly, the encompassing scope of our review, spanning diverse SDOH domains and emergency conditions, enriches the study’s comprehensiveness. Yet, the scope also poses challenges in distilling specific conclusions regarding the utility of data science techniques across distinct SDOH or EM conditions. Comparative analysis across studies was hampered by the varied outcome measures adopted. Although a significant number honed in on the performance of predictive models, this only scratches the surface of data science’s potential impact on SDOH within EM. Lastly, our review refrained from assessing the methodological quality of the incorporated studies. This approach aligns with scoping review guidelines but omits considerations of each study’s methodological soundness during our synthesis.

Amidst these intricacies and challenges, there are still other pressing concerns. Data quality, privacy concerns, and algorithmic biases are potential hurdles that merit attention. In particular, the limited exploration and assessment of algorithmic bias in our reviewed studies, given its potential to perpetuate healthcare disparities, suggests an urgent avenue for further investigation. Only one study we identified in our review specifically assessed ML model bias and fairness in the context of heart failure outcomes.^15^ ML algorithmic biases are critical to address in the context of SDOH research as prior studies have demonstrated the potential for reinforcement of pre-existing racial, ethnic, and socioeconomic disparities.^16^

### Future Works and Recommendations

#### Potential Areas of Exploration

Our review has illuminated the significance of certain domains like housing insecurity within the context of SDOH in Emergency Medicine. However, the vast landscape of SDOH offers numerous other domains that remain relatively unexplored:

1. Education: Investigating the role of educational attainment and access to quality education can provide insights into its impact on health outcomes. For instance, understanding how literacy levels influence patient adherence to medical advice in emergency settings could be pivotal.
2. Employment: Employment status, job security, and workplace conditions can have profound effects on mental and physical health. Exploring these factors can shed light on stress-related conditions or injuries that present in emergency departments.
3. Social Networks: The influence of social support systems, community engagement, and familial ties can play a crucial role in patient recovery and mental well-being. Delving into these aspects can offer a holistic view of a patient’s environment and its implications on health.

With these potential areas in mind, it becomes evident that a multi-faceted approach to SDOH within Emergency Medicine is the way forward. Building on these areas of exploration, we propose several recommendations to harness the full potential of SDOH in Emergency Medicine.

#### Recommendations

1. Establishing Gold Standard Metrics: For the evolution and standardization of emergency SDOH research, it is essential to define and adopt gold standard metrics. These metrics should be robust, universally accepted, and tailored to capture the nuances of SDOH in emergency settings. Collaborative efforts among researchers, clinicians, and policymakers should be made to create these benchmarks.
2. Innovative Data Capture: The high-paced nature of emergency settings necessitates innovative solutions for capturing SDOH data. Leveraging AI-assisted tools or predictive algorithms based on existing patient data could offer one approach.
3. Algorithmic Innovation: The prominence of machine learning and NLP in our findings suggests a horizon brimming with algorithmic advancements and adaptation for Emergency Medicine. As these tools evolve, and new tools emerge, crafting and evaluating interventions tailored to specific SDOH is crucial.
4. Connecting SDOH with Clinical Outcomes: Beyond identifying SDOH, understanding their tangible impact on patient outcomes is vital. A concerted effort in this direction can revolutionize our care approach.
5. Interdisciplinary Collaboration: The confluence of expertise, from clinicians to data scientists, will be instrumental in harnessing the full potential of SDOH data.
6. Addressing Algorithmic Bias: As we increasingly rely on algorithms, it is imperative to ensure they are free from biases that could perpetuate or exacerbate health disparities. Rigorous testing, validation, and refinement of algorithms, with a focus on fairness and equity, should be prioritized.

### Conclusion

This scoping review underscores the transformative potential of data science in elevating the understanding and application of Social Determinants of Health (SDOH) within Emergency Medicine. Through the adept integration of data science methodologies, particularly machine learning and natural language processing, we are poised to redefine the way SDOH data is adopted within EM. This offers a broader and more data-informed approach to influencing critical patient outcomes. The literature landscape indicates a promising embrace of this cross-disciplinary synergy, manifesting in an increasing number of studies that deploy data science methodologies to unearth, interpret, or model SDOH within emergency care contexts. Such a trajectory not only affirms the growing acknowledgment of these methodologies’ efficacy but also underlines the healthcare sector’s commitment to delivering more holistic care.

Nevertheless, our review also pinpoints avenues that warrant deeper exploration. Despite the expansive focus on various SDOH domains, certain determinants like housing insecurity and the neighborhood environment have garnered disproportionate attention. A more balanced exploration across SDOH domains would provide a richer, more comprehensive insight into their collective and individual impacts on patient trajectories. Moreover, while the current trend leans heavily on machine learning and natural language processing, there exists a vast expanse of data science techniques yet to be fully leveraged like large language modeling (LLMs). Diving into these untapped methodologies might further refine our capabilities in SDOH identification and intervention.

In conclusion, the fusion of data science with Emergency Medicine marks the dawn of a new healthcare epoch. It envisions a future where Emergency Departments transcend their traditional roles, evolving into hubs that address the foundational SDOH challenges within communities. As we navigate this promising trajectory, the potential to revolutionize EM and fortify patient-centric care is immense.

## Data Availability

All data produced in the present study are available upon reasonable request to the authors

## Acknowledgements

none

---

**Supplemental Table 1:** Finalized search strategies.

Embase Classic+Embase
|  |  |
| --- | --- |
| 1 | social determinants of health/ |
| 2 | ("Social determinants of health" or "Social risk" or "social need*").mp. |
| 3 | 1 or 2 |
| 4 | natural language processing/ |
| 5 | (Natural language processing or NLP).mp. [mp=title, abstract, heading word, drug trade name, original title, device manufacturer, drug manufacturer, device trade name, keyword heading word, floating subheading word, candidate term word] |
| 6 | machine learning/ |
| 7 | machine learning.mp. |
| 8 | artificial intelligence/ |
| 9 | ("artificial intelligence" or AI).mp. |
| 10 | clinical decision support system/ |
| 11 | Clinical decision support system*.mp. |
| 12 | Precision Medicine.mp. |
| 13 | predictive analytics.mp. |
| 14 | predictive model*.mp. |
| 15 | m-health.mp. |
| 16 | or/4-15 |
| 17 | emergency medicine/ |
| 18 | hospital emergency service/ |
| 19 | ("emergency medicine" or "emergency department*" or "emergency room*" or "emergency unit*" or "emergency ward*" or "emergency service*").mp. |
| 20 | 17 or 18 or 19 |
| 21 | 3 and 16 and 20 |

Cochrane CENTRAL Search
| ID | Search |
| --- | --- |
| #1 | MeSH descriptor: [Social Determinants of Health] this term only |
| #2 | ("Social determinants of health" OR "Social risk" OR "social need*"):ti,ab,kw |
| #3 | MeSH descriptor: [Natural Language Processing] this term only |
| #4 | ("Natural language processing" OR NLP):ti,ab,kw |
| #5 | MeSH descriptor: [Machine Learning] this term only |
| #6 | (machine learning):ti,ab,kw |
| #7 | MeSH descriptor: [Artificial Intelligence] this term only |
| #8 | ("artificial intelligence" OR AI):ti,ab,kw |
| #9 | MeSH descriptor: [Decision Support Systems, Clinical] this term only |
| #10 | ("Clinical decision support system*"):ti,ab,kw |
| #11 | MeSH descriptor: [Precision Medicine] this term only |
| #12 | (precision medicine):ti,ab,kw |
| #13 | ("predictive analytics"):ti,ab,kw |
| #14 | ("predictive model*"):ti,ab,kw |
| #15 | (m-health):ti,ab,kw |
| #16 | MeSH descriptor: [Emergency Medicine] this term only |
| #17 | MeSH descriptor: [Emergency Service, Hospital] this term only |
| #18 | (emergency medicine OR emergency department* OR emergency room* OR emergency unit* OR emergency ward* OR emergency service*):ti,ab,kw |
| #19 | {OR #1-#2} |
| #20 | {OR #3-#15} |
| #21 | {OR #16-#18} |
| #22 | #19 AND #20 AND #21 |

**Supplemental Table 2:** Database of Publications Meeting Final Inclusion Criteria.

| No. | Title | Authors | Pub. Year | Study Objective | Methodology | Models Utilized | Outcomes | ML Techniques | NLP Techniques | ML Tasks | NLP Tasks | SDOH Domains |
| --- | --- | --- | --- | --- | --- | --- | --- | --- | --- | --- | --- | --- |
| 1 | Inclusion of social determinants of health improves sepsis readmission prediction models | Amrollahi F, Shashikum ar SP, Meier A, Ohno-Machado L, Nemati S, Wardi G | 2022 | Deep learning model to predict 30-day readmissions in hospitalized sepsis patients incorporating clinical and large number of SDOH features. | Retrospective multicenter cohort study using medical and pharmacy claims data and neighborhood-level social determinants data | Neural network model with social determinants data compared to baseline models without social determinants data | Incorporating social determinants data into sepsis readmission prediction model improved AUC from 0.75 to 0.80 (p<0.001) | Neural network, Logistic regression | N/A | Readmission prediction | N/A | Multiple Domains (SES, education, healthcare access, ADL's, neighborhood and built environment) |
| 2 | Information Extraction From Electronic Health Records to Predict Readmission Following Acute Myocardial Infarction: Does Natural Language Processing Using Clinical Notes Improve Prediction of Readmission? | Brown JR, Rickett IM, Reeves RM, Shah RU, Goodrich CA, Gobbel G, Stabler ME, Perkins AM, Minter F, Cox KC, Dorn C, Denton J, Bray BE, Gouripeddi R, Higgins J, Chapman WW, MacKenzie T, Matheny ME | 2022 | NLP model to extract seven key social risk factors from clinical documentation to improve prediction 30-day readmission following acute myocardial infarction compared to structured clinical data model. | Linked data from EHRs and claims data; NLP used to extract social risk factors from clinical notes | Machine learning models with structured EHR data compared to models with added NLP-derived social risk factors | Adding NLP-derived social risk factors did not improve model performance for predicting 30-day readmissions after AMI | Elastic net, LASSO, Ridge regression, Random forest, Gradient boosting | Rule-based NLP system (Moonstone) | Readmission prediction | Entity extraction, attribute detection, document classification | Multiple Domains (social isolation/living alone, medication compliance/access, impaired ADL) |
| 3 | ReHouSED: A novel measurement of Veteran housing stability using natural language processing | Chapman AB, Jones A, Kelley AT, Jones B, Gawron L, Montgomery AE, Byrne T, Suo Y, Cook J, Pettey W, Peterson K, Jones M, Nelson R. | 2021 | NLP extraction and aggregation of clinical documentation for measurement of housing stability among a cohort of US Veterans. | Retrospective cohort study extracting housing-related information from clinical notes using NLP | Compared NLP-based measurement of housing stability to structured EHR data | NLP-based measure showed improved detection of housing instability compared to diagnosis codes | Rule-based NLP system | Entity extraction, attribute detection, document classification, patient-level housing status inference | N/A | Housing status classification, patient-level housing status inference | Housing Insecurity/homelessness |
| 4 | Neighborhood-level Social Determinants of Health Improve Prediction of Preventable Hospitalization and Emergency Department Visits | Chi W, Andreyeva E, Zhang Y, Kaushal R, Haynes K. | 2021 | Gradient boosting machine (GBM) modeling to determine if neighborhood level SDOH | Retrospective analysis using medical/pharmacy claims data linked to census block group-level social | Logistic regression models with claims data compared to models | Adding social determinants improved model performance for predicting preventable ED visits but | Logistic regression | N/A | Predicting preventable hospitalization and ED visits | N/A | Multiple Domains |
|  | Beyond Claims History |  |  | data could improve prediction of preventable hospitalization and ED visits. SDOH data improved prediction beyond using only claims data. | determinants data | with added neighborhood-level social determinants | not preventable hospitalizations |  |  |  |  |  |
| 5 | Development of a homelessness risk screening tool for emergency department patients | Doran KM, Johns E, Zuiderveen S, Shinn M, Dinan K, Schretzman M, Gelberg L, Culhane D, Shelley D, Mijanovich T | 2022 | Comparison of multiple ML modeling methods to develop a homelessness risk screening tool for emergency department patients to improve prediction for entering a homeless shelter. | Prospective cohort study using linked ED patient survey data and administrative homeless shelter data | Predictive modeling to identify risk factors for shelter entry after ED visit | 3-item screening tool identified risk for future homelessness with high sensitivity | Logistic regression, Classification and regression trees | N/A | Predicting risk of homelessness | N/A | Housing Insecurity/homelessness |
| 6 | A Statistical-Learning Model for Unplanned 7-Day Readmission in Pediatrics | Ehwerhemuepha L, Pugh K, Grant A, Taraman S, Chang A, Rakovski C, Feaster W. | 2020 | Employing least absolute shrinkage and selection operator (LASSO) regression the authors to examine risk of pediatric 7-day readmissions identifying 8 key features that modified risk of readmission. | Retrospective analysis of inpatient and observation encounters from a single pediatric hospital between 2013-2017. 50% of data used for model training, 50% for evaluation. | Least absolute shrinkage and selection operator (LASSO) regression | History of readmissions, medications, diagnoses, length of stay, and severity of illness were most significant predictors of 7-day readmissions. AUC 0.778. | LASSO regression | N/A | Classification, risk prediction | N/A | Multiple Domains |
| 7 | Your neighborhood matters: A machine-learning approach to the geospatial and social determinants of health in 9-1-1 activated chest pain | Faramand Z, Alrawashdeh M, Helman S, Bouzid Z, Martin-Gill C, Callaway C, Al-Zaiti S. | 2021 | Multiple ML methods were employed to examine interactions of geospatial factors and SDOH features to examine the likelihood of acute coronary | Developed random forest and logistic regression models to identify predictors of acute coronary syndrome using demographics, clinical data, | Random forest, regularized logistic regression | Residential neighborhood was a significant independent predictor of acute coronary syndrome, even after controlling for demographic | Random forest, logistic regression | N/A | Classification, feature importance | N/A | Neighborhood/Built Environment |
|  |  |  |  | syndrome among EMS attended chest pain calls. | geospatial coordinates, and census data. |  | s and clinical factors. |  |  |  |  |  |
| 8 | Using Clinical Notes and Natural Language Processing for Automated HIV Risk Assessment. | Feller DJ, Zucker J, Yin MT, Gordon P, Elhadad N. | 2018 | NLP and random forest classifiers were used to extract text and develop prediction models for an HIV risk assessment from clinical noted and electronic health records. The study employed social and behavioral determinants to improve prediction for identification of high-risk behavior. | Compared performance of models predicting HIV diagnosis using structured EHR data alone vs. with extracted clinical text topics vs. with extracted clinical text keywords. | Random forest classifier, regularized logistic regression | NLP improved model performance, especially using extracted keywords related to HIV risk factors. NLP keyword model achieved highest precision and recall. | Random forest, logistic regression | N/A | Classification, risk prediction | N/A | multiple domains (more behavioral and HIV risk factors) |
| 9 | Use of Stratified Cascade Learning to predict hospitalization risk with only socioeconomic factors. | Filikov A, Pethe S, Kelley R, Fischer A, Ozminkowski R. | 2020 | Use of a stratified cascade learning (SCL) model using only socioeconomic factors to predict risk of hospitalization among a cohort of diabetic patients. | Developed novel Stratified Cascade Learning framework to predict hospitalization risk using only socioeconomic and demographic variables. | Stratified Cascade Learning with random forest classification | The framework improved accuracy and specificity but reduced sensitivity compared to a baseline model. May help identify low-risk individuals. | Random forest | TF-IDF weighting, topic modeling with LDA, keyword identification with chi-square tests | Classification | Text preprocessing, keyword extraction, topic modeling | Multiple domains (mix of social and behavioral including insurance, access to care, smoking and ETOH) |
| 10 | Predicting opioid use disorder and associated risk factors in a Medicaid managed care population. | Gao W, Leighton C, Chen Y, Jones J, Mistry P | 2021 | Multiple machine learning methods were employed to develop and validate a predictive model for opioid use disorder (OUD) and prediction of future OUD among Medicaid patients. An indicator for SDOH vulnerability was included in the models. | Validation Study | Machine learning models (multivariate logistic regression, decision trees, random forest, neural networks, support vector machine) | Compared models and identified predictors for predicting opioid use disorder | Multivariate logistic regression, decision trees, random forest, neural networks, support vector machine | N/A | Prediction | N/A | Opioid Use Disorder |
| 11 | Predicting frequent emergency department use among children with epilepsy: A retrospective cohort study using electronic health data from 2 centers | Grinspan ZM, Patel AD, Hafeez B, Abramson EL, Kern LM | 2018 | Using multiple ML algorithms (Random Forest, adaBoost, support vector machines, and Lasso logistic regression), the authors developed a prediction model to for future ED usage among pediatric epilepsy patients. | Retrospective cohort study using EHR data from 2013 to predict ED use in 2014 at 2 pediatric centers | Logistic regression models compared to machine learning models like random forest, adaboost, support vector machine | 3-variable logistic regression model with prior ED use, insurance status, number of AEDs predicted ED use as well as machine learning models | logistic regression, random forest | n/a | classification, prediction | n/a | Multiple Domains, Subdomain: Area level SDOH (education, race, SES) |
| 12 | Multiple Electronic Health Record-Based Measures of Social Determinants of Health to Predict Return to the Emergency Department Following Discharge | Hall AG, Davlyatov GK, Orewa GN, Mehta TS, Feldman SS | 2022 | Development of predictive model employing weighted logistic regression and utilizing individual level SDOH and area deprivation index (ADI) to predict return to the ED within 30 days of hospital discharge. | Logistic regression analysis of EHR data from 2017-2018 to predict 30-day readmissions | Logistic regression models | Collectively SDOH variables made a relatively small contribution in determining readmission likelihood compared to medical conditions count | logistic regression | n/a | classification, prediction | n/a | Multiple Domains (both individual level including insurance status, working and living conditions, health status and disability, and area level measures including ADI) subcategory: area level |
| 13 | Classifying social determinants of health from unstructured electronic health records using deep learning-based natural language processing | Han S, Zhang RF, Shi L, Richie R, Liu H, Tseng A, Quan W, Ryan N, Brent D, Tsui FR | 2022 | Trained and evaluated three classes of deep neural networks (DNNs) for extraction of SDOH information from ICU database comparing to traditional | Annotated sentences from EHR notes for SDOH categories, developed and tested CNN, LSTM, BERT models | CNN, LSTM, BERT neural networks compared to logistic regression and random forests on bags of words | BERT model performed best overall with micro-F1=0.69, macro-AUC=0.907 | Convolutional neural network (CNN), long short-term memory (LSTM) network, Bidirectional Encoder Representations from Transformers (BERT) | N/A | Multi-label text classification | N/A | Multiple Domains (all major domains, behavioral determinants) |
|  |  |  |  | NLP techniques. |  |  |  |  |  |  |  |  |
| 14 | Community-based participatory research application of an artificial intelligence-enhanced electrocardiogram for cardiovascular disease screening: A FAITH! Trial ancillary study | Harmon DM, Adedinsewo D, Van't Hof JR, Johnson M, Hayes SN, Lopez-Jimenez F, Jones C, Attia ZI, Friedman PA, Patten CA, Cooper LA, Brewer LC | 2022 | Development of a community cardiovascular screening tool using AI-enhanced electrocardiograms for the detection of left ventricular ejection fraction for prediction and early detection of high-risk cardiovascular disease among an under-resourced cohort. | Validated AI-ECG model for age, sex, LVEF prediction in community-based cohort | Previously developed AI-ECG model | AI-ECG demonstrated good predictive performance for decreased LVEF and sex in the cohort |  |  |  |  | Area level measures (HOUSES index and ADI) |
| 15 | Development and assessment of a natural language processing model to identify residential instability in electronic health records' unstructured data: a comparison of 3 integrated healthcare delivery systems | Hatef E, Rouhizadeh M, Nau C, Xie F, Rouillard C, Abu-Nasser M, Padilla A, Lyons LJ, Kharrazi H, Weiner JP, Roblin D | 2022 | Validation of NLP algorithms for extraction of unstructured text in electronic health records for identification of homelessness and housing insecurity. | Developed and tested NLP algorithm to extract residential instability markers from EHR unstructured data at 3 healthcare systems | Rule-based NLP system using spaCy and compared to cTAKES | NLP algorithm performed with moderate precision, sensitivity and specificity at the 3 sites | N/A | Rule-based NLP using regular expressions | N/A | Information extraction, named entity recognition | Housing Instability/homelessness |
| 16 | Measuring the Value of a Practical Text Mining Approach to Identify Patients With Housing Issues in the Free-Text Notes in Electronic Health Record: Findings of a Retrospective Cohort Study | Hatef E, Singh Deol G, Rouhizadeh M, Li A, Eibensteiner K, Monsen CB, Bratslaver R, Senese M, Kharrazi H | 2021 | Advanced text mining strategies were employed to assess predictive value for identifying patients with homelessness and housing instability issues from electronic health record data. | Retrospective analysis of EHR data from a medical group using text mining (pattern matching with regex) to identify phrases related to housing issues | Rule-based text mining using regex patterns | High precision but low recall of the text mining approach compared to manual annotation | N/A | Pattern matching using regular expressions | N/A | Information extraction | Housing Instability/homelessness |
| 17 | Improving Fairness in the Prediction of Heart Failure Length | Li Y, Wang H, Luo Y | 2022 | Integrating key SDOH features and | Analysis of registry data to develop ML | Logistic regression, random | SDOH integration improved | Naive Bayes, logistic regression, | N/A | Binary classification, | N/A | Multiple domains: Area level SDOH (SDI and ADI) |
|  | of Stay and Mortality by Integrating Social Determinants of Health |  |  | area level indices (social deprivation index and area deprivation index), the study utilized 5 machine learning classifiers to predict hospital length of stay and in-hospital mortality among heart failure patients. | models predicting heart failure outcomes; assessed model fairness and effect of integrating SDOH data | forest, SVM, gradient boosting, naive Bayes | model fairness without compromising performance | support vector machine, random forest, gradient boosted decision trees |  | predicting probability scores |  |  |
| 18 | Developing a Model to Predict High Health Care Utilization Among Patients in a New York City Safety Net System | Li Z, Gogia S, Tatem KS, Cooke C, Singer J, Chokshi DA, Newton-Dame R | 2022 | Employing classification and regression tree (CART) and least absolute shrinkage and selection operator (LASSO) and key SDOH features, the study sought to predict health care utilization New York City Safety net hospital system. | Analysis of EHR data to develop model predicting high acute care utilization; incorporated social risk factors | Logistic regression, CART, LASSO | LASSO model with 17 predictors achieved highest PPV | Logistic regression, classification and regression tree (CART), least absolute shrinkage and selection operator (LASSO) | N/A | Classification, regression | N/A | Housing Instability/homelessness |
| 19 | Using Natural Language Processing to Examine Social Determinants of Health in Prehospital Pediatric Encounters and Associations with EMS Transport Decisions | Lowery B, D'Acunto S, Crowe RP, Fische JN | 2022 | NLP algorithms to extract SDOH data from free-text EMS registry data to quantify the association of SDOH features with EMS pediatric transport decisions. | Analysis of EMS records using NLP to extract SDOH factors and assess association with transport decisions | NLP using regex patterns | SDOH rarely documented but associated with higher transport odds when present | Logistic regression | Dictionary-based information extraction, preprocessing (lowercase, replace acronyms, correct misspellings, sentence boundary detection, lemmatization, negation detection) | Classification | Named entity recognition | Multiple domains (housing, food insecurity, insurance status, social support) |
| 20 | Methods for development and | Rousseau JF, Oliveira | 2022 | Development of an |  |  |  | N/A | N/A | N/A | N/A |  |
|  | application of data standards in an ontology-driven information model for measuring, managing, and computing social determinants of health for individuals, households, and communities evaluated through an example of asthma. | E, Tierney WM, Khurshid A. |  | ontological framework for improved integration of SDOH data among a pediatric asthma study population. |  |  |  |  |  |  |  |  |
| 21 | Identification of social determinants of health using multi-label classification of electronic health record clinical notes. | Stemerman R, Arguello J, Brice J, Krishnamurthy A, Houston M, Kitzmiller R. | 2022 | Multi-label learning to measure the performance for classification of SDOH from unstructured EHR data among mental health and substance use disorder patients utilizing the ED. | Retrospective analysis using multi-label classification models on EHR data | Linear SVM, KNN, Random Forest, XGBoost, Bi-LSTM | Identification of social determinants of health like financial strain and poor social support from clinical notes | Linear SVM, KNN, Random Forest, XGBoost, LSTM | Dictionary-based information extraction, preprocessing (tokenization, remove punctuation, lemmatization, stopword removal) | Multi-label classification | Named entity recognition | multiple domains (all major domains as defined by IOM) |
| 22 | Screening for Social Determinants of Health: Active and Passive Information Retrieval Methods. | Stewart de Ramirez S, Shallat J, McClure K, Foulger R, Barenblat L. | 2022 | A comparison and measurement of the performance of NLP algorithms on unstructured clinical text (passive) and active screening for identifying unmet SDOH needs among a tertiary referral hospital system. | Retrospective analysis comparing active vs passive screening | NLP models | Screening for social determinants using natural language processing | N/A | Dictionary-based information extraction | N/A | Named entity recognition | Multiple Domains (Economic, healthcare access/concerns, social isolation and physical risk) |
| 23 | Natural language processing and machine learning of electronic health records for prediction of first-time suicide attempts. | Tsui FR, Shi L, Ruiz V, Ryan ND, Biernesser C, Iyengar S, Walsh CG, Brent DA. | 2022 | NLP and ML algorithms were employed to both unstructured and structured data to predict first time suicide attempts. | Retrospective case-control study using NLP and ML on EHR data | Naive Bayes, LASSO, Random Forest, XGBoost | Prediction of first-time suicide attempts | Naive Bayes, LASSO, Random Forest, XGBoost | cTAKES for concept extraction and negation detection | Classification | Named entity recognition, negation detection | Multiple Domains (46+ features!!) |
| 24 | Prediction of emergency department revisits using area-level social determinants of health measures and health information exchange information. | Vest JR, Ben-Assuli O. | 2019 | Comparison of Two-Class Boosted of decision Tress ML algorithms to compare different data classes (EHR and HIE) and area level SDOH measures for the prediction of ED revisits. | Retrospective analysis comparing various data sources | Boosted decision trees | Prediction of emergency department revisits using social determinants and health information exchange data | Two-Class Boosted Decision Trees | N/A | Risk prediction modeling for ED revisits | N/A | Multiple Domains, Subcategory: Area level (US Census) |
| 25 | Measuring Exposure to Incarceration Using the Electronic Health Record. | Wang EA, Long JB, McGinnis KA, Wang KH, Wildeman CJ, Kim C, Bucklen KB, Fiellin DA, Bates J, Brandt C, Justice AC. | 2019 | Comparison of self-reported incarceration history across multiple data sources and NLP algorithms to unstructured text to identify exposure to incarceration among US Veteran's Administration patients. | Validation study using multiple data sources (self-report, DOC records, CMS records, VA records, NLP) | Linkage models, service code identification, NLP and machine learning | Compared different methods of identifying incarceration history in EHR to self-report. NLP had sensitivity of 63.5% and specificity of 95.9%. | N/A | Chart review, keyword search, Yale cTAKES extension, support vector machine classifier | N/A | Information extraction of incarceration exposure from EHR notes | Incarceration exposure |
| 26 | Comparison between machine learning methods for mortality prediction for sepsis patients with different social determinants. | Wang H, Li Y, Naidech A, Luo Y. | 2022 | After identifying disparities in key SDOH factors, machine learning classifiers were trained to predict in-hospital mortality among a cohort of sepsis patients. | Retrospective study using MIMIC-III database. Machine learning models trained to predict sepsis mortality. | 16 ML models tested: SVC, perceptron, random forest, ridge classifier, etc. | Statistically significant decreases in model performance for mortality prediction seen in Asian, Hispanic, and Spanish-speaking subgroups. | Ridge classifier, perceptron, passive-aggressive classifier, k-nearest neighbors (kNN), random forest, support vector machine with linear kernel (linearSVC) and L1 or L2 regularization, stochastic gradient descent (SGD) classifier with L1, L2, or elastic net regularization, multinomial naïve Bayes,<br>Bernoulli<br>naïve Bayes,<br>logistic<br>regression,<br>support<br>vector<br>machine<br>(SVM) with<br>rbf,<br>polynomial,<br>or sigmoid<br>kernel | N/A | Mortality prediction for sepsis patients | N/A | Multiple domains (marital status, insurance status, primary language, race) |

